# The impact of a women-led community center on social cohesion in Tripoli, North Lebanon: A pilot randomized controlled trial

**DOI:** 10.1101/2025.10.23.25338663

**Authors:** Maia Sieverding, Ahmad Addam, Maya Al Farran

**Affiliations:** American University of Beirut, Beirut, Lebanon; Threads of Peace, Beirut, Lebanon; Independent consultant, Salamanca, Spain

**Keywords:** Social cohesion, peacebuilding, forced displacement, civic engagement, wellbeing

## Abstract

**Background:** Social cohesion is essential to sustainable peace. Despite the global rise of armed conflict, evidence on effective approaches to building social cohesion in conflict-affected contexts is limited. This study evaluates the activities of a women-led community center on social cohesion among Lebanese and Syrian women in Lebanon, a country characterized by multiple displacement crises and deep sectarian divides.

**Methods:** Following an extensive recruitment drive in neighborhoods surrounding the community center, 206 women were randomized into an intervention and control group. The intervention group was invited to participate in seven sessions of training and structured dialogue facilitated by women volunteers at the center. A total of 177 women across the two groups completed both a baseline and endline survey capturing dimensions of social cohesion including trust, belonging and friendship networks, as well as civic engagement, wellbeing and self-efficacy. We present descriptive analysis of participants’ baseline characteristics, intervention take-up and social cohesion measures. We then conduct an intent-to-treat analysis of the intervention’s impact using regression models.

**Results:** Among women assigned to the intervention group, 70 (76.1%) attended any of the sessions. The intervention had a positive effect on participants’ sense of belonging, measured by the social relationship scale, and civic engagement attitudes and behaviors. We did not find effects on generalized trust or the other outcome measures.

**Conclusions:** This pilot randomized trial demonstrates the potential of community centers to foster aspects of social cohesion in fragile settings as well as the limitations of short, low-intensity contact interventions. The results emphasize the need for further investment in and evaluation of sustained, contextually adapted approaches. The study also shows that randomized evaluations of community-based initiatives are feasible in conflict-affected contexts and can generate valuable evidence to inform peacebuilding efforts.

## 1. Introduction

A quarter of the global population—approximately 2.1 billion people—live in fragile contexts, including 72% of those in extreme poverty [1]. The 2024 Global Peace Index reveals an historic decline in global peacefulness, with a record 97 countries becoming less peaceful, 56 active conflicts and a sharp rise in military spending and displacement [2]. As of June 2024, more than 37 million people were forcibly displaced across borders as refugees and 68 million more were displaced within their own countries due to conflict, persecution and disaster [3]. These trends highlight the urgent need for stronger investment in peacebuilding and conflict prevention over militarization [2].

In fragile contexts, such as those affected by conflict and forced displacement, social cohesion is critical to fostering peaceful coexistence, reducing inter- and intragroup tensions and supporting pathways to integration. This is particularly true where state structures are weakened or exclusionary [4,5]. Social cohesion, grounded in trust, a sense of belonging, participation, willingness to help and acceptance of diversity, offers a practical framework for mitigating social tensions and rebuilding fractured societies [4]. In a world increasingly shaped by uncertainty and instability, the question of how to effectively build social cohesion is critical to promoting peace and supporting sustainable post-conflict and post-disaster reconstruction. However, rigorous evidence on how to build social cohesion in fragile contexts is limited; a recent systematic review identified only 37 papers based on 24 unique studies across all Low- and Middle-Income Countries (LMICs) [4]. This paper contributes to this limited evidence base by reporting the results of a pilot randomized controlled trial (RCT) of a locally developed, community-based intervention that aimed to promote social cohesion in Lebanon, a fragile state hosting large Syrian and Palestinian refugee populations and affected by deep sectarian divisions and armed conflict.

### 1.1 Social cohesion

There is no single, universally accepted definition of social cohesion; rather, scholars have articulated the concept in multiple ways, reflecting diverse theoretical frameworks and contextual interpretations. Generally, social cohesion refers to the strength and quality of relationships that bind individuals and groups within a society, enabling cooperation, trust, and a shared sense of belonging. It encompasses both horizontal ties, i.e. the connections between individuals and communities, and vertical ties that link people to institutions and the state [6]. The literature broadly agrees on five interrelated domains of social cohesion: trust, sense of belonging, willingness to participate, willingness to help and acceptance of diversity [4,7].

Social cohesion is strongly linked to sustainable peace. Sustainable peace involves fostering meaningful connections among diverse social groups, promoting inclusive participation in governance processes and ensuring that traditionally marginalized groups have a voice in decision-making [8].

Evidence from both high-income countries and LMICs shows that the most successful social cohesion interventions are those that address both structural inequalities and inter-group relations [4,7,9,10]. For example, in contexts affected by displacement, policies that ensure equitable access to basic services, early inclusion of host communities in planning processes and joint economic development opportunities reduce exclusion and foster shared interests [11–13]. Infrastructure such as inclusive public spaces and shared service facilities play a vital role in building trust and a sense of belonging [14]. However, such policies and infrastructure are often lacking in LMIC contexts shaped by internal divisions and where political approaches to displaced populations are often exclusionary [15].

At the individual and communal level, interventions rooted in contact theory, which involves fostering positive, cooperative interactions between members of different social or identity groups, ideally under conditions of equality and a supportive institutional environment, have generally shown small positive effects on social cohesion [4]. These interventions include modalities such as sports teams, collaborative projects and facilitated dialogues that bring people from different groups together [4]. In the Southwest Asia and North Africa (SWANA) region, contact theory-based interventions implemented among conflict-affected communities have yielded mixed results. In Iraq, a mixed-religion soccer intervention demonstrated positive impacts on Christian players’ attitudes towards Muslim teammates but not on inter-group cohesion more broadly [16]. In Lebanon, participation in a national youth volunteering program had a positive impact on Lebanese youths’ sense of belonging and tolerance [17]. However, two other training-based interventions found, respectively, no impact on inter-group prejudice between Lebanese and Syrian refugees, and potentially some backlash among Lebanese participants [18], and no impact on Jordanian and Lebanese host communities’ outgroup bias but some small positive impacts among Syrian refugees [19]. Explanations for these divergent findings may include variation in program design and the short duration of most interventions, the different groups involved (in the context of Lebanon’s multiple social cleavages), and the different measures used to capture social cohesion.

### 1.2 Community center approaches

Interventions grounded in contact theory highlight the importance of structured opportunities for positive, cooperative interaction between groups. Community centers represent a promising but underexplored modality for operationalizing this approach in fragile and refugee-hosting contexts. By providing shared spaces that facilitate equal access, repeated interaction and collective activities, they offer an institutionalized infrastructure for fostering neighborhood-level social cohesion, particularly in marginalized urban areas [20].

Evidence from high-income settings demonstrates the role of community spaces in strengthening social ties and rebuilding fragmented social fabrics. The urban planning literature emphasizes that “space matters”: the provision of open and inclusive community spaces can increase social interaction, build social capital and enhance sustainability [20]. Similar findings have been reported in regeneration projects in Europe, including community center models in Switzerland designed to enhance participation and belonging in diverse neighborhoods [21] and in more recent work linking community centers to social entrepreneurship and collaborative innovation [22].

Community centers have also shown promise in displacement contexts. Refugee resettlement programs in the United States highlight the role of such spaces in creating cross-group engagement and supporting integration [21]. In the SWANA region, programming for Syrian refugees that provided spaces that facilitate cross-group engagement and shared service access [23,24], thereby operating in ways comparable to community centers, also suggests the potential of this approach to build sense of belonging and stronger inter-group ties.

Despite the promise of community centers as participatory, community-led spaces for building social cohesion, they remain largely absent from the literature in LMICs and are rarely subject to evaluation. To the best of our knowledge, no previous studies in the SWANA region have evaluated a community center-based approach to building social cohesion. The only systematic review of social cohesion interventions in LMICs to date also did not identify any such evaluations [4]. Given their adaptability and grounding in participatory, community-led approaches, this highlights an imperative gap in the literature regarding effective approaches for fostering social cohesion and stability in fragile contexts.

## 2. Methods

### 2.1. Setting

Tripoli, Lebanon’s second-largest city and the capital of the Northern Governorate, has long grappled with structural marginalization, deteriorating infrastructure and chronic underinvestment. These conditions have been exacerbated by Lebanon’s ongoing economic collapse since 2019, with poverty rates in Tripoli exceeding 50% and unemployment surpassing 35% in some estimates [25,26]. The city also hosts large numbers of Syrian and Palestinian refugees, concentrated in impoverished urban neighborhoods. The resulting pressure on basic services and economic opportunities has intensified existing socio-economic vulnerabilities and intergroup tensions between host and refugee populations [25,26]. Within this fragile urban landscape, this study focuses on two adjacent but deeply divided neighborhoods: Bab al-Tabbaneh (predominantly Sunni) and Jabal Mohsen (predominantly Alawite), separated by the symbolic and contested Syria Street. These neighborhoods are nationally emblematic of entrenched sectarian fault lines and have experienced what scholars have described as “the deadliest proxy war in Lebanon” [27]. Between these two neighborhoods, long-standing tensions escalated with the spillover of the Syrian conflict, culminating in recurrent rounds of armed clashes between 2008 and 2015 that left dozens dead, injured hundreds, and displaced families on both sides [28,29]. Despite modest improvements in local relations and apparent signs of reduced clashes, Bab al-Tabbaneh and Jabal Mohsen remain heavily stigmatized and continue to stand as potent reminders of the fragility of Lebanon’s social cohesion and the lingering volatility of urban communal relations [29].

### 2.2. The women-led community center in Tripoli and the evaluated activities

Situated between the divided neighborhoods of Bab al-Tabbaneh and Jabal Mohsen, the women-led community center known as Beit el Salam was established in 2021 as a neutral and inclusive space for dialogue, collaboration, and peacebuilding. The center was created with the support of Threads of Peace (commonly known as House of Peace, or HOPe), a local peace-building NGO in Lebanon. HOPe is a politically neutral, non-sectarian organization whose core model, the Social Peace Process, brings together community members from diverse backgrounds to engage in dialogue, build trust, jointly analyze challenges and design initiatives that address real needs. The process aims to empower participants as leaders of change and fosters resilience, inclusion and social cohesion within their communities.

The establishment of Beit Al Salam center resulted from the coming together of four previously independent, women-led initiatives established through HOPe’s Social Peace Process. These initiatives consist of two cooking groups, an upcycling project and a wellness care initiative. Although each had distinct goals, the idea of a shared space emerged as both a practical solution—allowing them to lower operational costs—and a symbolic act of unity. In its early stages, the center operated primarily as a shared platform through which the initiatives continued their own activities, but in a collective and more visible way. The simple act of women from both communities working side by side and being seen together in public service became a symbol of cooperation across divides. Over time, the center has evolved into a broader hub for women-led activities and local engagement. While the women themselves lead and manage these efforts, HOPe provides technical support.

The intervention evaluated in this study marked the first structured, multi-session program to be designed and delivered entirely by women from the center. Developed in collaboration with HOPe, the intervention brought together participants into fixed groups that met weekly over the course of seven sessions of about two and a half hours each. As a pilot currently under evaluation for potential scale-up, the program combined trainings by the center’s four initiatives with structured dialogue and group-building components. Each session was grounded in a shared agreement to ensure a safe, respectful environment, and opened with a group check-in to foster emotional connection. Sessions were facilitated by the volunteer women who had previously launched the community-led initiatives that gave rise to the center itself—an intentional effort to deepen their leadership and sustain their role as agents of change within their community. Women were not compensated for attending the sessions. Transportation costs to the community center were not covered by the program, but transportation was facilitated through a shared bus service with group pick-up points in the neighborhoods.

### 2.3. Study design

The study was implemented as a randomized controlled trial (RCT). The RCT protocol was registered on the American Economic Association RCT Registry (ID: AEARCTR-0013423). Community center volunteers conducted an intensive outreach campaign to recruit women for a series of sessions to be held at the center. They distributed digital flyers via the center’s social media platforms that provided information on the center’s activities and a link for interested women to fill a registration form. Volunteers also conducted in-person recruitment in neighborhoods, filling the recruitment form for those who expressed interest in the activities. The recruitment form collected contact and basic demographic information on potential participants, including information needed to screen for eligibility. The eligibility criteria for participation were that the individual be a woman, aged 18 or above, of Lebanese, Palestinian or Syrian nationality, living in the target communities and who had not previously attended activities at the community center. Recruitment was conducted between April 29, 2024 and July 31, 2024.

A total of 522 unique individuals filled out the recruitment form. After applying eligibility criteria, 362 women were identified as eligible to participate (Figure 1). These individuals were contacted via WhatsApp by trained women volunteers from the community center, who conducted an initial welcoming call. During the call, volunteers explained the program’s objectives, the nature of participation, and logistical considerations such as the absence of compensation or transportation coverage. Following these calls, 206 women confirmed their interest in joining the sessions and agreed to be contacted regarding the evaluation study. This was considerably below our initial target sample size of 340 participants, based on sample size calculations using monitoring data that HOPe had collected from other workshops, albeit with different populations. Transportation emerged as a significant challenge to participation; although the program subsequently facilitated a shared bus with group pick-up points, it remained a barrier for many eligible participants who lived too far from the pick-up locations to join. The center’s initiatives were also concerned about their ability to offer the sessions to such a large number of women given space constraints.

**Fig 1:**
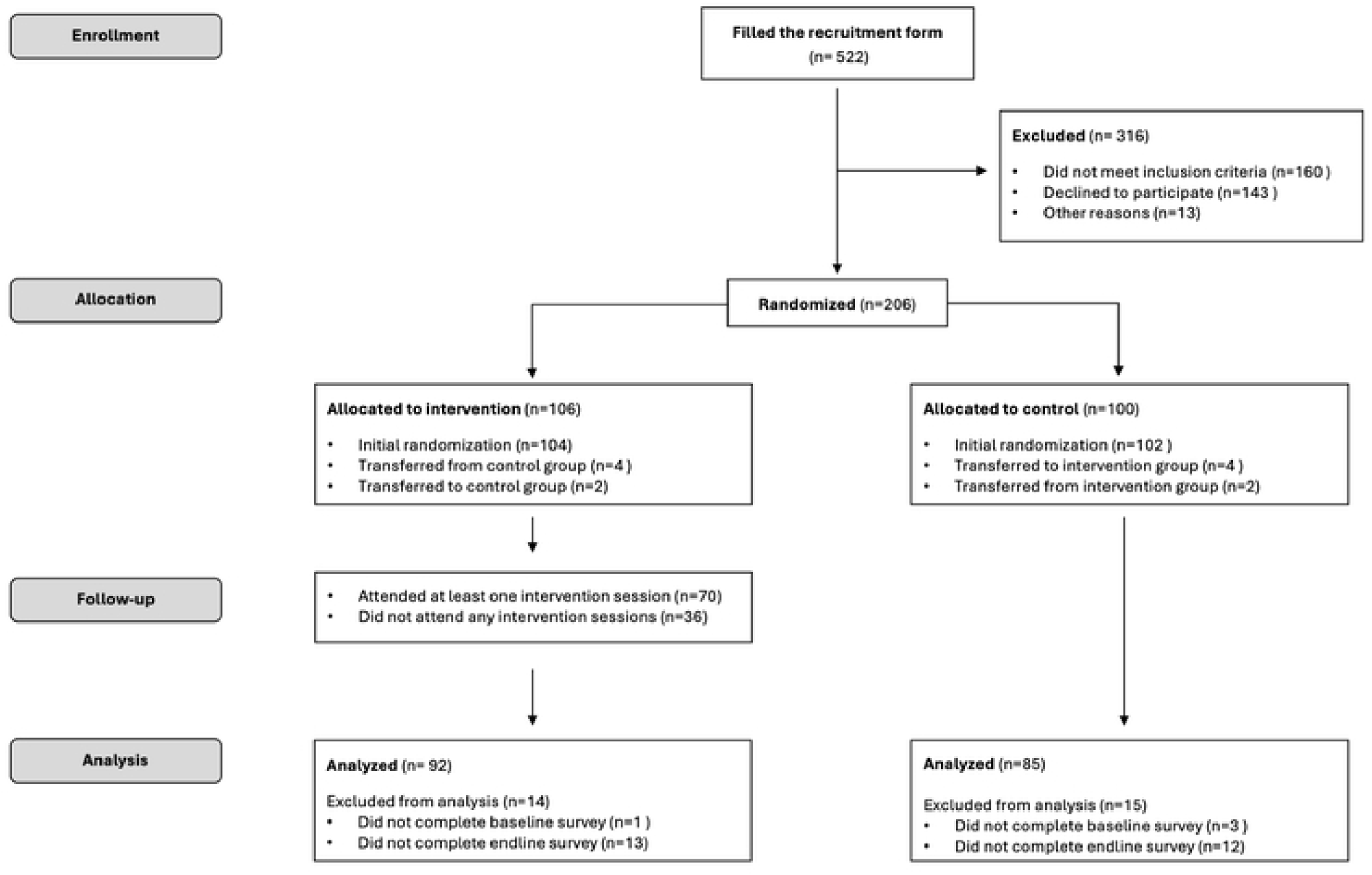
Study flowchart.

Randomization was conducted using Stata by taking a simple random sample, without replacement, of 50% of the de-identified participant list. A small number of participants who registered late were randomized separately using the same process. Women who were selected in the sample were assigned to the intervention group that was invited to participate in the community center trainings. Women who were not selected were assigned to a control group that was informed they would be offered a space in the training sessions in the fall of 2024. Due to delays related to the war in Lebanon in fall 2024, women in the control group were offered the intervention sessions in spring 2025. The randomization results were communicated to the participants after the baseline survey was completed so as not to bias survey responses.

Mobility is a challenge for women in the study context. After randomization was completed, several participants contacted the implementation team to inform them that they could only attend the sessions if they could commute to the center with a friend or relative who had been randomized to the other group. To avoid losing women from the center and the study, these requests were accommodated. Four women were therefore moved from their original control group assignment to the intervention group and two women from the intervention group to the control group (Figure 1).

### 2.4. Data collection

Data collection was conducted at baseline (pre-intervention) and endline (post-intervention) using structured, self-administered surveys completed via an online form. Based on HOPe’s previous experience and given the sensitive nature of the topic, self-administered questionnaires were preferred to reduce response bias based on social desirability and (perceived) characteristics of the interviewer. The surveys captured respondents’ basic sociodemographic information and outcomes related to social cohesion, civic engagement, self-efficacy and subjective wellbeing. Questions on intervention attendance and satisfaction were added for the intervention group at endline.

All recruited participants were contacted by the evaluation team by phone prior to being informed of the results of the randomization. Using an approved recruitment script and consent form, female undergraduate student volunteers from a local university obtained oral informed consent to participate in the study, emphasizing that participation was voluntary, confidential, and would not affect participants’ involvement in the program. The volunteers documented that consent was obtained on an Excel-based study tracking sheet. Consenting participants were then sent the baseline survey link via WhatsApp. While the survey was self-administered, participants with limited literacy or digital access were supported by the same student volunteers, who conducted the survey verbally and recorded responses on their behalf following standardized protocols. A total of 202 participants completed the baseline survey between June and July 2024 (Figure 1). All surveys were collected in the Levantine dialect of Arabic on the KoBo Toolbox platform.

One month following completion of the intervention, a link to the endline survey was sent to all study participants via a WhatsApp to assess short-term outcomes. A total of 177 participants completed the endline survey between October and November 2024 (Figure 1; 12.4% attrition). There was no evidence of selective attrition. Allocation to the intervention vs. control group was not significantly associated with study attrition, nor was nationality or neighborhood of residence (Supplemental Table A1). Of collected sociodemographic characteristics, only younger age was significantly associated with attrition. Given these results and the fact that the endline data collection was conducted during the 2024 war on Lebanon, attrition appears to have been largely random and related to the difficult circumstances in the country.

### 2.5. Outcomes

The study examined outcomes in three areas: social cohesion, civic engagement and wellbeing. To the extent possible, we employed validated outcome measures from existing literature. Other measures were developed or adapted to the context given the paucity of related measures that had been developed and tested in a Middle Eastern context. Due to the sensitivity of the topic, “prefer not to answer” options were added for all outcome questions. These responses were coded to missing at the analysis stage, leading to incomplete data for some outcomes.

Our primary outcome was generalized trust, one of the core components of social cohesion. We measured trust through a single-item question used, among other studies, in the global World Values Survey program: “Generally speaking, would you say that most people can be trusted or that you need to be very careful when dealing with people?” [30,31]. The option choices were “Most people can be trusted,” “You need to be very careful” and “Don’t know.” The small number of “Don’t know” (n=6 at endline) responses were recoded as “You need to be very careful” for analysis. Additionally, a three-item generalized trust scale was constructed using questions about whether the respondent trusts 1) people she meets for the first time, 2) people from different religions and 3) people from different nationalities. Responses were collected on a 4-point Likert scale (1 = *Do not trust at all, 2 = Do not trust very much, 3 = Trust somewhat,* 4 = *Trust completely)* and averaged across the three items to yield a continuous score between one and four, with higher values indicating greater trust.

We also used the Social Relationships Scale developed by Wilson and Secker [32] in the United States. The scale was chosen to capture the sense of belonging dimension of social cohesion. The eight-item scale assesses perceived inclusion, safety, and social integration in the community. Items captured respondents’ perceived usefulness to the community, feelings of being appreciated by others, geographic mobility and cultural exposure, social and cultural participation, and experiencing prejudice and safety concerns. Responses to each item were recorded on a 4-point scale ranging from *Not at all*=1 to *Yes, definitely*=4. We added a “prefer not to say” option that was recoded to missing at the analysis stage. To ensure consistent interpretation across items, we reverse coded negatively phrased items at the analysis stage so that higher values always indicate more positive perceptions of belonging. We then calculated a continuous, composite score by summing all items into a scale with a raw score range from 8 – 32. The reliability of the scale was acceptable, although not optimal, with an alpha of 0.71. To faciliate interpretation, we standardized the scale to have a mean of zero and a standard deviation of one so that the results can be interpreted in terms of change in standard deviations.

Our other measure of social cohesion assessed diversity of social ties. Respondents were asked how many close female friends they had and, of those friends, how many were of a different religion and how many were of a different nationality. As responses were clustered at the lower end of the distribution, we created a categorical outcome for each question coded as 0 friends, 1–2 friends, 3– 4 friends and 5 or more friends.

We measured civic engagement, which overlaps with the social cohesion domain of willingness to help, using the Civic Engagement Scale developed by Doolittle and Faul [33] in the United States. The scale captures both attitudinal commitments—such as social responsibility and civic duty—and behavioral expressions of helping, volunteering, and community participation. The tool includes two subscales measuring civic attitudes and behaviors, respectively. The attitudinal subscale was comprised of eight items reflecting participants’ beliefs about social responsibility, civic duty, volunteering and involvement in the community. Responses were recorded on a 7-point Likert scale (*Disagree*=1 to *Agree*=7), with higher scores indicating stronger civic values. We calculated a continuous civic attitudes score by summing all eight items (scale range 7-56). The subscale had acceptable reliability in the study population with an alpha of 0.79. The behavioral subscale included five of the original six items in the subscale, which assesses actual civic actions such as volunteering, helping others, donating to charity and engaging in community discussions. Responses were similarly recorded on a 7-point scale and summed to create a civic engagement score (range 5-35). The subscale also had acceptable reliability with an alpha of 0.76. We then derived a total civic engagement score by summing the attitudinal and behavioral subscales (range 12-91). Alpha of the overall scale was 0.82. We standardized all three scales to have a mean of zero and a standard deviation of one.

Our measure of subjective wellbeing was the WHO-5, a short, simple tool with demonstrated validity across a wide range of contexts and interventions [34]. The WHO-5 captures participants’ self-reported positive emotional states over the previous two weeks. Respondents rate the frequency of experiencing five states, such as “I have felt cheerful and in good spirits” on a 6-point scale from 0 (“*At no time”*) to 5 (“*All of the time”*). The five items were summed (range 0–25) and multiplied by four to yield a final score ranging from 0 to 100, with higher scores indicating better well-being [34].

Finally, we measured generalized self-efficacy (GSE), which captures an individual’s global belief in her ability to manage new or difficult tasks or to cope with adversity [35]. To facilitate respondent understanding of the self-efficacy questions, we adopted a 3-item General Self-Efficacy Scale (GSE-3) scale [36] despite the fact that this scale has not been previously well-validated in Arabic (unlike the longer, original GSE scale). The GSE-3 consists of three statements “I can rely on my own abilities in difficult situations,” “I am able to solve most problems on my own,” and “I can usually solve even challenging and complex tasks well.” Response choices are on a 5-point Likert scale ranging from 1 (“*Does not apply at all*”) to 5 (“*Applies completely*”). Responses across the three items were averaged to create a single, continuous measure with a range from one to five [36]. Alpha of the scale was 0.72.

### 2.6. Analysis

We present descriptive analysis of the characteristics of the intervention and control groups at baseline, demonstrating a balanced evaluation sample. We also present descriptive analysis of compliance with the randomized assignment, focusing on attendance of the community center sessions among women who were assigned to the intervention group. Given the lack of evidence on social cohesion measures among our study population in general, we then present a descriptive analysis of the outcome measures at baseline. For all descriptive analyses, statistical significance tests are conducted using a chi-squared test for categorical variables and a t-test for continuous variables.

Our estimate of the intervention impact is based on an intent-to-treat (ITT) analysis, i.e. we consider women’s assignment in the RCT regardless of whether they complied with the assignment. This approach is appropriate for an intervention, such as the one under study, in which compliance was imperfect and take-up of the intervention is key to its success [37]. In other words, whether people are willing to commit to an uncompensated, community-based program is an important aspect of whether such programs can be effective and sustainable. Thanks to the randomized design, the ITT estimates are straightforward and entail a simple regression of the outcome on treatment status.

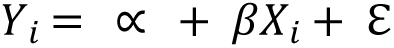

Where *Y_i_* is the outcome of interest for individual Y and X is equal to 1 if the individual was in the intervention group and 0 otherwise. For continuous outcomes we estimate and Ordinary Least Squares (OLS) regression model. For binary outcomes, we estimate a logistic regression model and for ordinal outcomes (number of friends) an ordinal logistic regression model.

### 2.7. Ethical approval

Ethical approval for the study was obtained from the American University of Beirut Institutional Review Board (Protocol SBS-2023-0360).

## 3. Results

### 3.1. Characteristics of participants

The randomization was successful, with no significant differences in the sociodemographic characteristics of the intervention and control groups (Table 1). Participants were on average 38.5 years old and around two-thirds were between ages 35 and 64. Two-thirds of participants were Lebanese and the remainder were Syrian. No significant differences were found in the religious makeup of neighborhoods where participants resided (p=0.23), although the percentage of women in the control group who lived in mixed neighborhoods was somewhat higher. Overall, 28.6% of participants lived in primarily Sunni neighborhoods, 25.6% in primarily Alawite neighborhoods and 45.7% in mixed neighborhoods, reflecting the success of the recruitment strategy in reaching women from different communities.

**Table 1:** Balance between intervention and control groups at baseline (all observations at <u>baseline)</u>.

|  | Control<br>N=97 | Intervention<br>N=105 | Total<br>N=202 | p-value |
| --- | --- | --- | --- | --- |
| **Age (categorical)** |  |  |  | 0.25 |
| 18-34 | 33 (34.4%) | 35 (33.3%) | 68 (33.8%) |  |
| 35-64 | 63 (65.6%) | 67 (63.8%) | 130 (64.7%) |  |
| >=65 | 0 (0.0%) | 3 (2.9%) | 3 (1.5%) |  |
| **Age** | 38.9 (10.3) | 38.2 (11.2) | 38.5 (10.8) | 0.68 |
| **Nationality** |  |  |  | 0.99 |
| Lebanese | 62 (63.9%) | 67 (63.8%) | 129 (63.9%) |  |
| Syrian | 35 (36.1%) | 38 (36.2%) | 73 (36.1%) |  |
| **Neighborhood** |  |  |  | 0.23 |
| Primarily Sunni neighborhoods | 26 (27.4%) | 31 (29.8%) | 57 (28.6%) |  |
| Primarily Alawite neighborhoods | 20 (21.1%) | 31 (29.8%) | 51 (25.6%) |  |
| Mixed neighborhoods | 49 (51.6%) | 42 (40.4%) | 91 (45.7%) |  |
| **Marital status** |  |  |  | 0.82 |
| Married | 69 (72.6%) | 77 (74.0%) | 146 (73.4%) |  |
| Not married | 26 (27.4%) | 27 (26.0%) | 53 (26.6%) |  |
| **Employment status** |  |  |  | 0.27 |
| Employed | 16 (16.8%) | 14 (13.5%) | 30 (15.1%) |  |
| Unemployed | 42 (44.2%) | 58 (55.8%) | 100 (50.3%) |  |
| Not in labor force | 37 (38.9%) | 32 (30.8%) | 69 (34.7%) |  |
| **Heard of community center before** |  |  |  | 0.54 |
| Yes | 47 (49.5%) | 56 (53.8%) | 103 (51.8%) |  |
| No | 48 (50.5%) | 48 (46.2%) | 96 (48.2%) |  |

*Notes: p-values are based on a Pearson's chi-square test for categorical outcomes and a t-test for* *continuous outcomes.*

Three-quarters of participants were married and the large majority were not working. Just over half reported having heard of the community center prior to the recruitment drive for this intervention, again with no significant difference (p=0.54) between women allocated to the intervention and control groups. Balance on all assessed characteristics of the intervention and control groups was maintained at endline despite attrition from the study (Supplemental Table A2).

### 3.2. Take-up of the intervention

Of the 92 women in the intervention group with complete observations, 70 (76.1%), attended at least one of the 7 sessions (Table 2). This leaves a notable proportion, 22 (24%), who never attended any session (of the women in the intervention group who did not complete the endline survey, 9 of 12 also had not attended any intervention sessions). While participants had been informed prior to the start of the intervention that no compensation or transportation support would be provided, when contacted to ask why they did not attend the sessions difficulty commuting to the center was one of the more common reasons. Other reasons included personal or family health issues, caregiving responsibilities and return to Syria. No sociodemographic characteristics were associated with never attending the sessions, although the estimates are very imprecise due to the small sample size (Supplementary Table A3). Looking more closely at the session attendance, the data shows considerable variation in engagement levels among those who attended. Only about a third of women in the intervention group attended all sessions. On the other hand, among women assigned to the control group and who were succesfully surveyed at endline, 16 (20.0%) reported having visited the community center since the baseline survey was run. However, as the intervention sessions were closed and center volunteers monitored attendance, it is likely that these visits were for other reasons, e.g. open community events or social gatherings.

**Table 2:** Intervention take-up and compliance with treatment assignment.

| Intervention | Control |
| --- | --- |

|  | N=92 | N=85 |
| --- | --- | --- |
| **Ever attended** |  |  |
| Never attended | 22 (23.9%) | - |
| Ever attended | 70 (76.1%) | - |
| **Total sessions attended** |  |  |
| Never attended | 22 (23.9%) | - |
| Only 1 session | 4 (4.3%) | - |
| 2 sessions | 3 (3.3%) | - |
| 3 sessions | 11 (12.0%) | - |
| 4 sessions | 4 (4.3%) | - |
| 5 sessions | 8 (8.7%) | - |
| 6 sessions | 9 (9.8%) | - |
| All sessions | 31 (33.7%) | - |
| **Visited the center since baseline** |  |  |
| No | - | 66 (80.0%) |
| Yes | - | 16 (20.0%) |

**Table 3:** Descriptive analysis of intervention outcomes at baseline.

|  | Control<br>N=97 | Intervention<br>N=105 | Total<br>N=202 | p-value |
| --- | --- | --- | --- | --- |
| **Social cohesion** |  |  |  |  |
| **Most people can be trusted (%)** |  |  |  | 0.84 |
| Careful/DK | 84 (88.4%) | 91 (87.5%) | 175 (87.9%) |  |
| Yes | 11 (11.6%) | 13 (12.5%) | 24 (12.1%) |  |
| **Generalized trust (3-item) (mean, SD)** | 2.5 (0.7) | 2.4 (0.7) | 2.5 (0.7) | 0.87 |
| **Social relationship scale (mean, SD)** | 22.2 (5.1) | 21.3 (5.1) | 21.7 (5.1) | 0.31 |
| **Number of friends (categorical, %)** |  |  |  | 0.41 |
| 0 friends | 1 (1.1%) | 4 (3.9%) | 5 (2.6%) |  |
| 1-2 friends | 33 (37.1%) | 32 (31.4%) | 65 (34.0%) |  |
| 3-4 friends | 21 (23.6%) | 31 (30.4%) | 52 (27.2%) |  |
| 5+ friends | 34 (38.2%) | 35 (34.3%) | 69 (36.1%) |  |
| **Number of friends (mean, SD)** | 4.9 (5.0) | 5.0 (5.3) | 4.9 (5.2) | 0.90 |
| **No. friends diff religion (categorical, %)** |  |  |  | 0.45 |
| 0 friends | 37 (45.1%) | 50 (52.6%) | 87 (49.2%) |  |
| 1-2 friends | 29 (35.4%) | 26 (27.4%) | 55 (31.1%) |  |
| 3-4 friends | 8 (9.8%) | 13 (13.7%) | 21 (11.9%) |  |
| 5+ friends | 8 (9.8%) | 6 (6.3%) | 14 (7.9%) |  |
| **Friends of different religion (mean, SD)** | 1.7 (3.3) | 2.1 (3.8) | 1.9 (3.5) | 0.47 |
| **No. friends diff nationality (categorical, %)** |  |  |  | 0.78 |
| 0 friends | 33 (38.8%) | 44 (45.4%) | 77 (42.3%) |  |
| 1-2 friends | 33 (38.8%) | 36 (37.1%) | 69 (37.9%) |  |
| 3-4 friends | 13 (15.3%) | 11 (11.3%) | 24 (13.2%) |  |
| 5+ friends | 6 (7.1%) | 6 (6.2%) | 12 (6.6%) |  |
| **Friends of different nationality (mean, SD)** | 1.5 (2.2) | 1.7 (3.3) | 1.6 (2.8) | 0.70 |
| **Civic engagement** |  |  |  |  |
| **Civic engagement attitudes (mean, SD)** | 42.8 (10.5) | 44.2 (10.4) | 43.5 (10.5) | 0.36 |
| **Civic engagement behaviors (mean, SD)** | 21.3 (7.7) | 20.3 (8.5) | 20.8 (8.2) | 0.37 |
| **Civic engagement scale (mean, SD)** | 64.1 (16.0) | 64.5 (15.9) | 64.3 (15.9) | 0.89 |
| **Wellbeing** |  |  |  |  |
| **WHO-5 (mean, SD)** | 32.7 (24.7) | 34.0 (27.1) | 33.3 (25.9) | 0.73 |
| **GSE-3 (mean, SD)** | 3.5 (0.8) | 3.5 (0.8) | 3.5 (0.8) | 0.77 |

### 3.3. Social cohesion, civic participation and wellbeing among study participants

At baseline, participants showed a high level of social mistrust, with nearly 90% reporting that one must be too careful in dealing with people. Scores on the other measures were somewhat higher. On the three-item social trust scale, the mean score was 2.5 (out of a maximum of four). On the social relationship scale, the mean score was 21.7, somewhat above the middle on the possible range of 8 – 32. Nearly all respondents reported having close female friends, with a mean of 4.9. On average, of these approximately five close friends, 1.9 were of a different religion and 1.6 of a different nationality. However, nearly half (49.2%) had no friends of a different religion and 42.3% had no friends of a different nationality, indicating considerable variation in diversity of participants’ social networks.

Turning to civic engagement, participants scored on average 43.5 on the attitudes subscale, well above the midpoint, and 20.8 on the behaviors subscale, right at the midpoint, suggesting more positive attitudes towards civic engagement than realized behaviors. The average score on the overall scale was 64.3. The mean score on the WHO-5 was low, at 33.3, well below the midpoint of a scale with a maximum of 100. The average score on the GSE-3 was 3.5 out of five. There were no significant differences in any of the outcomes by treatment status, indicating balance between the intervention and control groups at baseline.

### 3.4. Impact of the intervention (ITT estimates)

Table 4 presents the estimates of the intervention impact on measures of social cohesion. Intervention participants had 12% higher odds of saying that in general people can be trusted, but the result was not statistically significant. There was no impact on participants’ reported number of friends of a different nationality or on the three-item social trust scale. There was an impact on the social relationship scale, with participants in the intervention group scoring 0.60 standard deviations higher than those in the control group at endline (p<0.001). However, there was also a substantial amount of missing data on this variable, indicating that some respondents did not want to answer the scale questions.

**Table 4:**
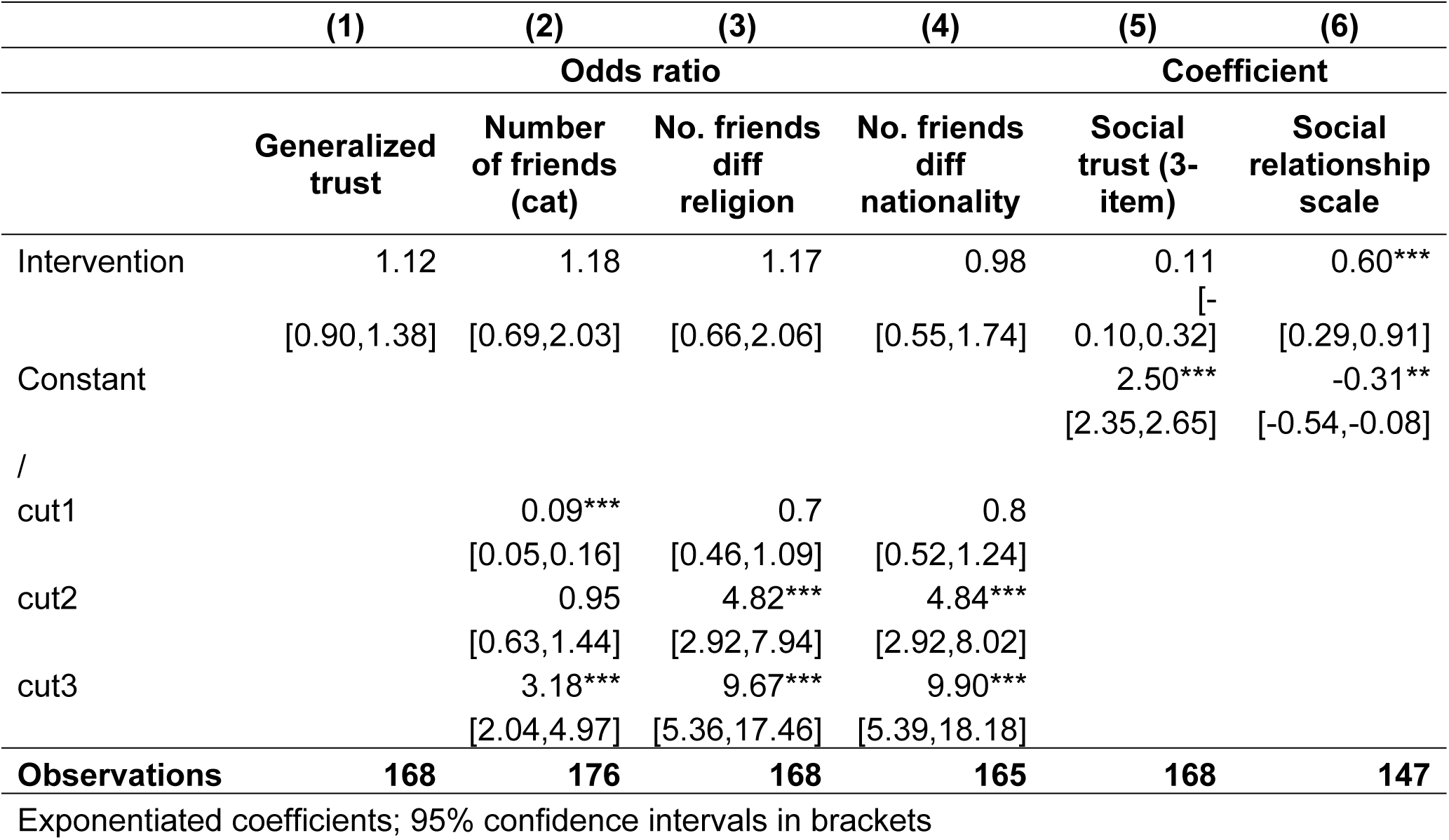
Intervention impacts on measures of social cohesion (logistic regression, ordinal logistic regression and OLS regression results)

Turning to the other outcome measures, the intervention had a significant impact on civic engagement attitudes and behaviors (Table 5). Participants in the intervention group scored 0.33 standard deviations higher on the attitudes subscale at endline (p<0.05) and 0.26 standard deviations higher on the behaviors subscale (p<0.1), for an increase of 0.33 standard deviations (p<0.05) on the overall scale. No significant impacts were observed on generalized self-efficacy or subjective wellbeing.

**Table 5:** Intervention impacts on measures of civic engagement, wellbeing and self-efficacy (OLS regression results)

|  | (1) | (2) | (3) | (4) | (5) |
| --- | --- | --- | --- | --- | --- |
|  | Civic<br>engagement<br>attitudes | Civic<br>engagement<br>behaviors | Civic<br>engagement<br>scale | GSE-3 | WHO-5 |
| **Intervention** | 0.33* | 0.26^ | 0.33* | 0.08 | -1.89 |
|  | [0.04,0.63] | [-0.04,0.55] | [0.04,0.63] | [-0.18,0.35] | [-9.50,5.71] |
| **Constant** | -0.17 | -0.13 | -0.17 | 3.52*** | 32.33*** |
|  | [-0.38,0.05] | [-0.34,0.08] | [-0.38,0.04] | [3.33,3.71] | [26.85,37.81] |
| **Observations** | **177** | **177** | **177** | **177** | **177** |

95% confidence intervals in brackets
^ p<0.1, \* p<0.05, \*\* p<0.01, \*\*\* p<0.001

## 4. Discussion

We report the results of a pilot randomized controlled trial of activities implemented by a women-led community center located between two conflict-affected neighborhoods in Tripoli, Lebanon. To our knowledge, this is the first RCT to evaluate a community center–based peacebuilding initiative in a fragile setting and in the SWANA region. In addition to generating robust evidence on whether participation in the center’s programming leads to measurable improvements in social cohesion outcomes, the study contributes to the literature by assessing the feasibility of implementing a randomized evaluation of a locally implemented initiative in a fragile and low-resource setting. Evaluating such local initiatives is essential to building the evidence base on what works to promote social cohesion in conflict-affected settings, particularly in the absence of strong state institutions or comprehensive national integration policies.

Through this evaluation, we found that participation in the sessions implemented by the community center had a positive impact on women’s sense of belonging, captured by the social relationships scale, and civic engagement attitudes and behaviors. We found no impact on generalized trust, number of close friends from out-groups, self-efficacy or subjective wellbeing. These results, and specifically those related to social cohesion outcomes, are broadly consistent with the literature on fairly low-dose, collaborative contact interventions, where more immediate effects are often seen in relational and participatory domains, whereas deeper or more global constructs such as generalized trust tend to require longer exposure or higher intensity to shift [4].

In the social cohesion literature, civic engagement is widely considered to encompass the “willingness to help” and “willingness to participate” domains, which together reflect an individual’s readiness to act for the betterment of the group through mutual support, volunteering and civic involvement [6,38]. Positioning our findings within this conceptual framework allows us to benchmark the study’s results against a broad body of evidence. The results are broadly consistent with global evidence that light-touch collaborative interventions can generate early shifts in participation and belonging such as rural community-driven development programs in Colombia that increased cooperative behaviors or framed collective-action exercises in Haiti that spurred volunteering [39].

Collaborative contact interventions in Iraq, Lebanon, and Nigeria similarly report small but significant improvements in sense of belonging and willingness to participate [16,17,40], with similar patterns observed for workshop-based interventions in Jordan and Nigeria, which also showed modest positive effects on belonging and participation [4]. It is worth noting that variation in how different aspects of social cohesion are measured is a major challenge in synthesizing this literature, and these studies used outcome measures that were different from those that we used [4]. However, taken together, these parallels suggest that the community center’s activities produced effects comparable in direction and magnitude to other rigorously evaluated interventions in fragile and divided settings.

Consistency with the broader literature on social cohesion interventions is not limited to the positive outcomes observed in this study. The broader literature on collaborative contact interventions has often found null or mixed results on trust when interventions are of short duration or relatively low intensity. For example, a collaborative contact intervention in Iraq similarly found no significant effect on generalized trust, although the intervention itself consisted of a soccer league among men [16], so both the population and intervention modality were distinct.

An explanation for the lack of impact on our primary outcome, trust, may be that short duration, relatively low-intensity collaborative contact interventions are not effective in shifting this outcome. Trust is a complex and deeply ingrained social behavior that may not shift significantly in just seven sessions. A longer intervention, with additional components or ongoing reinforcement could potentially lead to more measurable change [39]. Meta-analyses of workshop-based peace education interventions implemented over several years further support this interpretation. In a global synthesis, five such programs—most lasting four to five years and involving substantial geographic and participant coverage—were found to have a positive and significant effect on trust, suggesting that both duration and intensity play a critical role in producing measurable attitudinal change [4].

Another explanation for the lack of effect on trust and some of our other outcomes of interest may be the methodological challenges faced by the RCT. Although we had limited data on which to base initial power calculations, our ideal sample size was nearly twice the number of women we were able to enroll. The evaluation was thus likely under-powered to detect change in some outcomes. The small sample size also limited our ability to disaggregate results by participant characteristics, an important consideration given that prior research shows that the effectiveness of social cohesion interventions in Lebanon varies by nationality [18,19] and potentially by sect.

Equally important is what we measured and how. Our measures of social cohesion were largely drawn from validated scales, but scales that were developed in contexts very different to that of Lebanon, which may have constrained our ability to detect the most relevant and context-specific changes emerging from the program. For example, our global measure of self-efficacy was broad and may have missed domain-specific growth in areas such as conflict resolution skills, which participants covered during the training. The measure on close friendships may not have captured other, meaningful but not as deeply formed connections between participants. Unfortunately, it was beyond the scope of the study to engage in more extensive measures development. In general, the paucity of rigorously developed and tested measures of social cohesion in Middle Eastern contexts is a major gap in the literature on a region where conflict is so pervasive. Finally, the conditions under which data were collected are critical to interpreting the study’s results. The endline survey took place roughly one month after the intervention and coincided with a period of heightened political instability and active conflict. Many participants were coping with fear, economic uncertainty and disruption to daily life at the time of data collection that may have affected their responses.

These limitations notwithstanding, we believe that this study demonstrates that rigorous evaluation of locally implemented interventions can be feasible and makes an important contribution to the literature on how to build social cohesion in fragile contexts. The randomization of participants was successful and barriers to recruitment and adherence were largely related to transportation. Other challenges to randomization, such as women’s mobility constraints, could potentially be managed on a larger scale by randomizing women in pairs or small groups formed around commuting. Attrition from the study was within acceptable bounds, particularly given that the endline was conducted during the 2024 war under very challenging conditions. Online data collection with modalities to support participants with low literacy or visual impairment was critical in this regard.

Our results also demonstrate the potential for community center-based approaches to building social cohesion in fragile contexts. Despite the challenges with sample size, we found positive impacts on measures of sense of belonging and willingness to participate, consistent with literature on other interventions in other contexts. It is also important to emphasize that this study captures the effect of a specific, time-bound intervention within the community center, rather than the center’s broader role in shaping social cohesion at the community level. The women recruited for the RCT were secondary participants with respect to the center’s activities; the primary participants—the women who manage and deliver initiatives—who have deeper and more sustained engagement with diverse groups, could not be captured within the present evaluation design. Future work related to the potential impact of the community center modality will ideally look at sustainability, both in terms of the center’s operations and the effects of longer-term engagement with the center. Given the potential of intervention approaches that use intergroup contact in combination with economic support [4], strengthening the training aspects of the center’s initiatives in terms of their vocational training potential, in addition to serving as a platform for intergroup contact, is also a promising way forward in the difficult economic context of Lebanon.

## Data Availability

All relevant data for reproducing study outcomes are provided within the manuscript and its Supporting Information files. The data required to reproduce Table 1 are not provided in full because the combination of sociodemographic variables presented in the table could potentially allow for the identification of study participants.

## Appendix

**Table A1:** Correlates of attrition from the study.

|  |  |
| --- | --- |
| **Intervention group** | 1.1 |
|  | [0.43,2.78] |
| **Age** | 0.94* |
|  | [0.90,0.99] |
| **Did not previously know of center** | 0.45 |
|  | [0.15,1.32] |
| **Neighborhood (ref: Primarily Sunni)** |  |
| Primarily Alawite neighborhoods | 0.77 |
|  | [0.20,2.99] |
| Mixed neighborhoods | 0.65 |
|  | [0.21,1.95] |
| **Nationality (ref: Lebanese)** |  |
| Syrian | 0.87 |
|  | [0.30,2.53] |
| **Employment status (ref: employed)** |  |
| Unemployed | 4.57 |
|  | [0.54,38.55] |
| Not in Labor Force | 3.89 |
|  | [0.43,35.21] |
| **Not married** | 2.17 |
|  | [0.80,5.88] |
| Observations | 199 |

Exponentiated coefficients; 95% confidence intervals in brackets
\* p<0.05, \*\* p<0.01, \*\*\* p<0.001

**Table A2:**
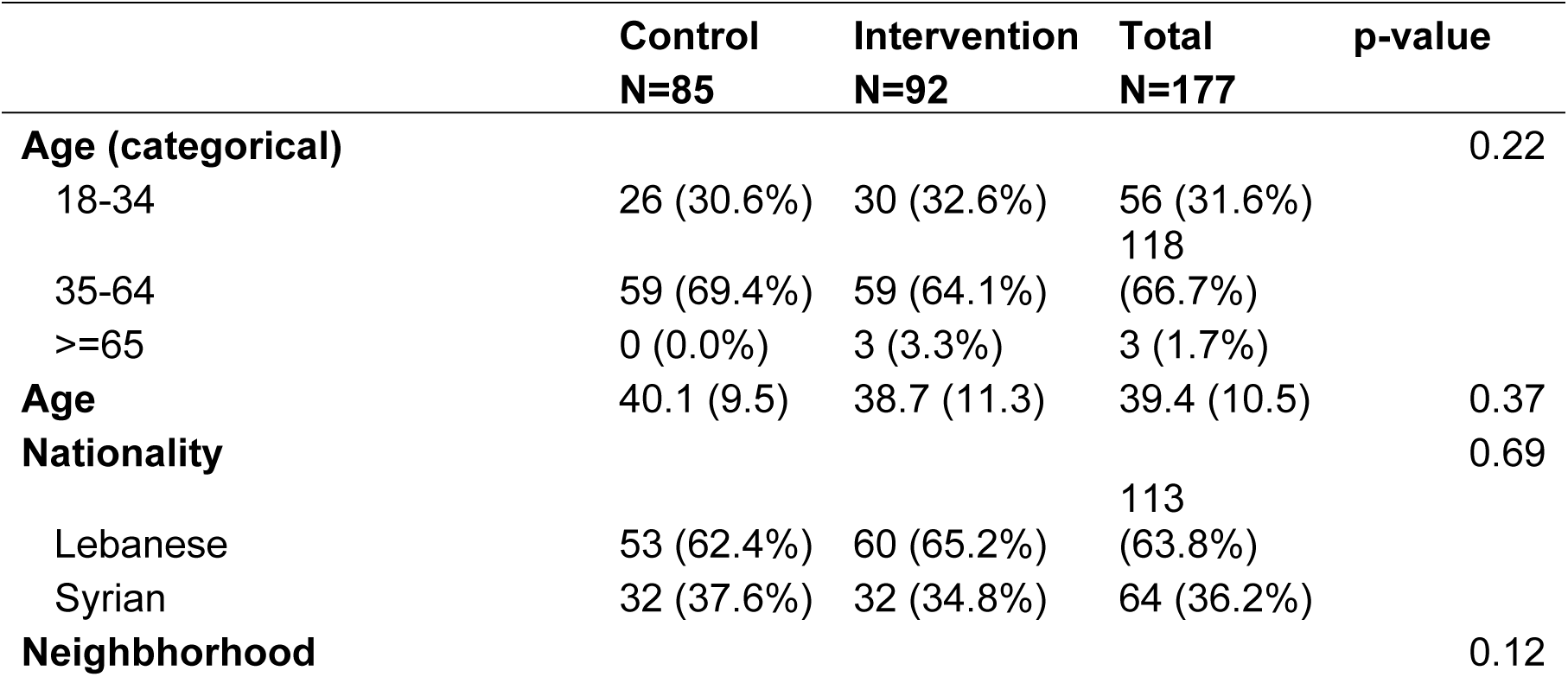
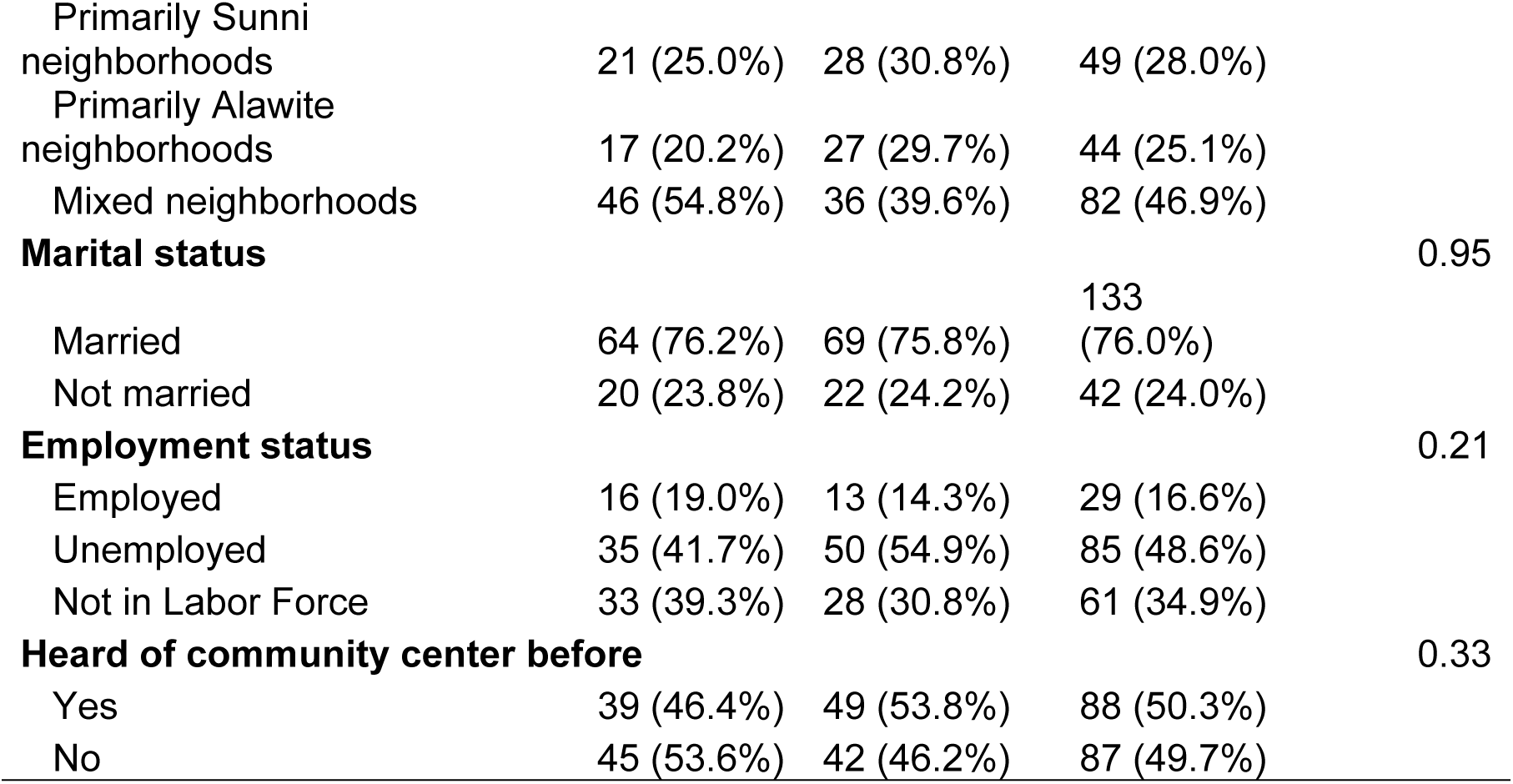
Balance between intervention and control groups at baseline (complete panel observations only)

**Table A3:** Correlates of never attending the intervention sessions (among women assigned to the intervention group)

|  |  |
| --- | --- |
| **Age** | 0.96 |
|  | [0.90,1.01] |
| **Did not previously know of center** | 0.55 |
|  | [0.15,2.00] |
| **Neighborhood (ref: Primarily Sunni)** |  |
| Primarily Alawite neighborhoods | 1.21 |
|  | [0.26,5.71] |
| Mixed neighborhoods | 1.12 |
|  | [0.31,4.10] |
| **Nationality (ref: Lebanese)** |  |
| Syrian | 1.04 |
|  | [0.30,3.53] |
| **Employment status (ref: employed)** |  |
| Unemployed | 3.47 |
|  | [0.62,19.50] |
| Not in Labor Force | 0.9 |
|  | [0.12,7.07] |
| **Not married** | 1.16 |
|  | [0.33,4.12] |
| Observations | 91 |

\* p<0.05, \*\* p<0.01, \*\*\* p<0.001

## Notes

### Competing Interest Statement

The authors have declared no competing interest.

### Clinical Trial

The RCT is not a clinical trial. The field of research is the behavioral and social sciences, so the trial was registered in the American Economics Association RCT registry (ID: AEARCTR-0013423).

